# LymphGen-Sig: Integrating genetic and transcriptional states to predict therapeutic response in DLBCL

**DOI:** 10.64898/2026.02.20.26346725

**Authors:** Sravya Tumuluru, Alan Cooper, Yanwen Jiang, Connie Lee Batlevi, Will Harris, Gilles Salles, Marek Trneny, Georg Lenz, Franck Morschhauser, Fabrice Jardin, Sandhya Balasubramanian, Matthew Sugidono, Alex F. Herrera, Justin Kline, James K. Godfrey

**Author notes:** Corresponding Author: James Godfrey, M.D., Department of Hematology & Hematopoietic Cell Transplantations, City of Hope 1500 East Duarte Road, Duarte, CA 91010. These authors contributed equally (Sravya Tumuluru, Alan Cooper, Justin Kline, James K. Godfrey).

## Abstract

**Purpose:** Genetic classification may advance precision medicine in diffuse large B-cell lymphoma (DLBCL), but existing tools like LymphGen (LG) are limited by complexity, incomplete classification, and do not incorporate non-genetic features that impact disease biology and therapeutic outcomes. To address these limitations, we developed LymphGen-sig (LGsig), a gene expression-based platform that classifies all DLBCLs and harmonizes both genetic and non-genetic dimensions of the disease.

**Methods:** LGsig was built on the distinct subtype-specific gene expression signature of each LG class using paired genomic and transcriptomic data (NCI/BCCA; n=764). Model development was restricted to DLBCLs classified into MCD, BN2, EZB, or ST2. Gene features were selected by differential gene expression, with 294 genes being optimal for classification using a nearest shrunken centroid classifier. LGsig classifications were designated *MCDsig*, *BN2sig*, *ST2sig*, and *EZBsig*. The final model was applied to RNAseq from archival samples from the POLARIX trial (n=678) to assess outcomes after polatuzumab vedotin-R-CHP (pola-R-CHP) or R-CHOP for each LGsig subtype.

**Results:** LGsig accurately identified LG subtypes using transcriptional data alone, and extended assignments to all previously LG-unclassified cases. Importantly, LG-unclassified DLBCLs reassigned by LGsig, mirrored the transcriptional and clinical features of their corresponding LG counterparts, supporting their reclassification. Additionally, LGsig reassigned LG A53 DLBCLs, characterized by aneuploidy and *TP53* alterations, into more biologically and therapeutically-relevant LGsig clusters. Lastly, LGsig improved the performance of LG as a biomarker in the POLARIX study, by identifying distinct DLBCL subtypes exhibiting a survival benefit with pola-R-CHP over R-CHOP in both LG-classified and LG-unclassified cases.

**Conclusion:** LGsig expands molecular classification beyond current genetic classifiers in DLBCL by integrating both genetic and transcriptional dimensions of the disease to better inform subtype-specific therapeutic strategies.

## Introduction

Diffuse large B-cell lymphoma (DLBCL) is the most common lymphoma diagnosed in the United States with approximately 25,000 new cases recognized each year.^1^ Chemoimmunotherapy regimens such as R-CHOP (Rituximab, Cyclophosphamide, Doxorubicin, Vincristine, and Prednisone) and polatuzumab vedotin (pola)-R-CHP achieve cure rates of roughly 60-70% in newly diagnosed patients.^2–4^ However, most patients who experience relapsed or refractory disease are not cured with available second-line therapies, and DLBCL remains responsible for more deaths than any other lymphoma subtype.^2,5–7^

In an effort to improve patient outcomes, numerous studies have sought to apply precision medicine approaches to DLBCL. Cell-of-origin (COO) has been the most widely used biomarker in precision medicine–based clinical trials.^8^ However, data from multiple large, randomized studies have indicated that COO alone is insufficient to guide therapeutic decisions.^9,10^ More recently, genomic classification systems have refined DLBCL classification beyond COO.^11–19^ Although several genetic classifications systems have been developed, the most widely adopted has been a probabilistic classifier called LymphGen (LG), which organizes DLBCLs into seven distinct subtypes based on co-occurring mutations, copy number alterations, and chromosomal translocations.^11^ Retrospective and prospective studies have shown that employing LG to inform treatment selection in the front-line setting can improve patient survival, establishing genomic classification as a promising platform for precision medicine in DLBCL.^20,21^

However, LG has important limitations, including its technical complexity and cost, which have hampered its clinical implementation. Furthermore, LG fails to classify approximately 40% of DLBCLs, leaving many patients without a clear subtype assignment for therapeutic guidance.^11^ In addition, genomic approaches such as LG fail to incorporate non-genetic determinants of DLBCL behavior and treatment response, such as transcriptional programs and microenvironmental features. These limitations highlight the need for complementary strategies that capture the functional consequences of genetic alterations and the broader biological context of the disease. Transcriptional profiling offers such an approach. For example, we recently developed a CD5 gene-expression signature (CD5sig) and demonstrated its ability to identify a subset of DLBCLs with enhanced sensitivity to Bruton tyrosine kinase inhibitor (BTKi)–based therapy in the phase III PHOENIX study.^22^ Notably, CD5sig captured DLBCLs with both genetic and non-genetic mechanisms of BTKi response, underscoring the value of transcriptional profiling for uncovering functional biology not revealed through genomic analysis alone.

Building on these insights, we sought to develop an integrated classification framework that reflects both the genetic and transcriptional dimensions of DLBCL. Using a LG transcriptional classifier, termed LymphGen-sig (LGsig), we demonstrate that each LG genetic subtype exhibits distinct and reproducible gene-expression features that enable accurate subtype prediction from bulk transcriptional data alone. Importantly, DLBCLs unclassified by the original LG algorithm share transcriptional and clinical features with canonical LG subtypes, allowing their reclassification into biologically related groups by LGsig and increasing the proportion of cases that can be meaningfully assigned. We also find that LGsig refines LG subgroups such as A53, revealing biological and clinical distinctions obscured by genomic data alone. Finally, application of LGsig to archival tumor samples from the randomized phase III POLARIX trial identifies DLBCL subtypes with superior progression-free survival (PFS) and overall survival (OS) following front-line treatment with pola-R-CHP compared to R-CHOP.^3,23^ Together, these findings establish LGsig as a robust and clinically relevant platform that bridges genetic classification and functional biology in DLBCL.

## Methods

### Data sets

#### NCI cohort

Data for 481 DLBCL samples were downloaded from dbGaP (phs001444), including RNA-seq counts, mutation calls, copy number alterations, and gene fusions.^11,24^

#### BCCA cohort

Data for 283 DLBCL samples were provided by BCCA or downloaded from previous publications, including RNA-seq counts, mutation calls, copy number alterations, and gene fusions.^14,25^

#### POLARIX

Data for 678 DLBCL samples were provided by Roche/Genentech, including RNA-seq counts and mutation calls from whole-exome sequencing.

### Gene expression analysis

#### RNA-seq processing

Normalization of RNA-seq counts was performed using a variance-stabilizing transformation (*vst*) in DESeq2 (v1.36.0).^26^ Where appropriate, batch correction was performed using the *removeBatchEffect* function in limma (v3.52.4).^27^

#### Differential gene expression (DGE)

DGE was performed using the limma *voom* method, using TMM (trimmed mean of M values)-normalization in the edgeR (v 3.40.2)^28^ and pre-filtering with a cutoff of 0.5 log2 counts-per-million.

#### Molecular classifications

Gene-expression-based cell of origin (COO) classifications for the NCI cohort were determined from their DGE-based classifier^24^ (Schmitz et al.), and COO classifications for the BCCA cohort were from Lymph2Cx.^29^ RNA-seq-based classifications for Dark Zone signature (DZsig) for NCI and BCCA samples were from Wright et al (there termed DHITsig), and PMBLsig classifications were used from Lymph3Cx.^30^ CD5sig was applied as previously described.^22^ B cell states were recovered in transcripts-per-million (TPM)-normalized data using the online Ecotyper tool (https://ecotyper.stanford.edu/lymphoma/).^18^

### Building LymphGen-Sig

#### Samples

In a combined cohort of NCI-BCCA, samples were restricted to those with LymphGen-core classifications of MCD, BN2, ST2, and EZB. A53 DLBCLs were excluded as a model class due to lack of a reproducible transcriptional signature. N1 DLBCLs were excluded due to limited case numbers. The remaining 276 samples were split 75%-25% stratified by subtype to define training and testing sets.

#### Feature selection

Genes enriched in a particular LymphGen subtype were determined using DGE analysis. DGE was performed comparing each subtype to the average of every other subtype (e.g., MCD vs (BN2 + ST2 + EZB)/3), and the top *n* genes meeting thresholds of adj. p < 0. 05 and |log2FC| > 1 were retained; duplicate genes found in multiple comparisons were condensed. The procedure was repeated for multiple *n* ranging from 200 genes to 600 genes in steps of 50.

#### Model development

The nearest shrunken centroid algorithm was selected for the model and was trained in a pipeline that included random oversampling of non-majority classes followed by feature scaling with z-score normalization. The shrinking threshold (range 0.01 to 1, steps of 0.01) was tuned using 5-fold stratified cross-validation scored using a macro-weighted F1-score. All algorithms were from scikit-learn (v1.3.2)^31^, and random oversampling was performed using imblearn (v0.11.0)^32^, both of which were run in Python (v3.9.17) via reticulate (v1.35.0).

### Statistical methods

All statistical tests were performed in R (v4.2.1). Statistical significance for categorical variables was assessed with a Fisher’s exact test for two-group comparisons and a chi-squared test for more than two groups. Survival analyses were performed using the survival package (v3.4.0), and the log-rank test was used to determine significance. A p-value threshold of 0.05 was used to assess univariate statistical significance. For POLARIX, Hazard ratios (HR) were adjusted for IPI score (2 vs 3–5), age (<60 vs ≥60 years) and COO. All statistics are descriptive. Adjustment for multiple hypothesis testing was performed using the Benjamini-Hochberg method.

## Results

### LG subtypes are accurately recapitulated by transcriptional features

To build the LGsig classifier, we first defined transcriptional features that distinguished conventional LG subtypes by performing differential gene expression (DGE) analyses on cohorts of diagnostic DLBCL specimens from the National Cancer Institute and British Columbia Cancer Agency (NCI/BCCA; n = 764).^11,14^ Here, we identified gene features associated with each LG subset (MCD, BN2, ST2, EZB) by comparing each group to all remaining LG-classified DLBCLs (e.g., MCD vs. non-MCD) (**Supplementary Figure 1A-D)**. The N1 subtype was excluded due to small sample size. In total, we identified 959 significantly differentially expressed genes (DEGs) distinguishing each LG subtype. To visualize the candidate transcriptional features identified through DGE analysis, we generated a heatmap of gene-wise z-scored expression across all samples for the top DEGs, **(Figure 1A)**. As expected, each LG subtype was associated with a specific gene expression program, suggesting that each is transcriptionally distinct from the others.

**Figure 1.**
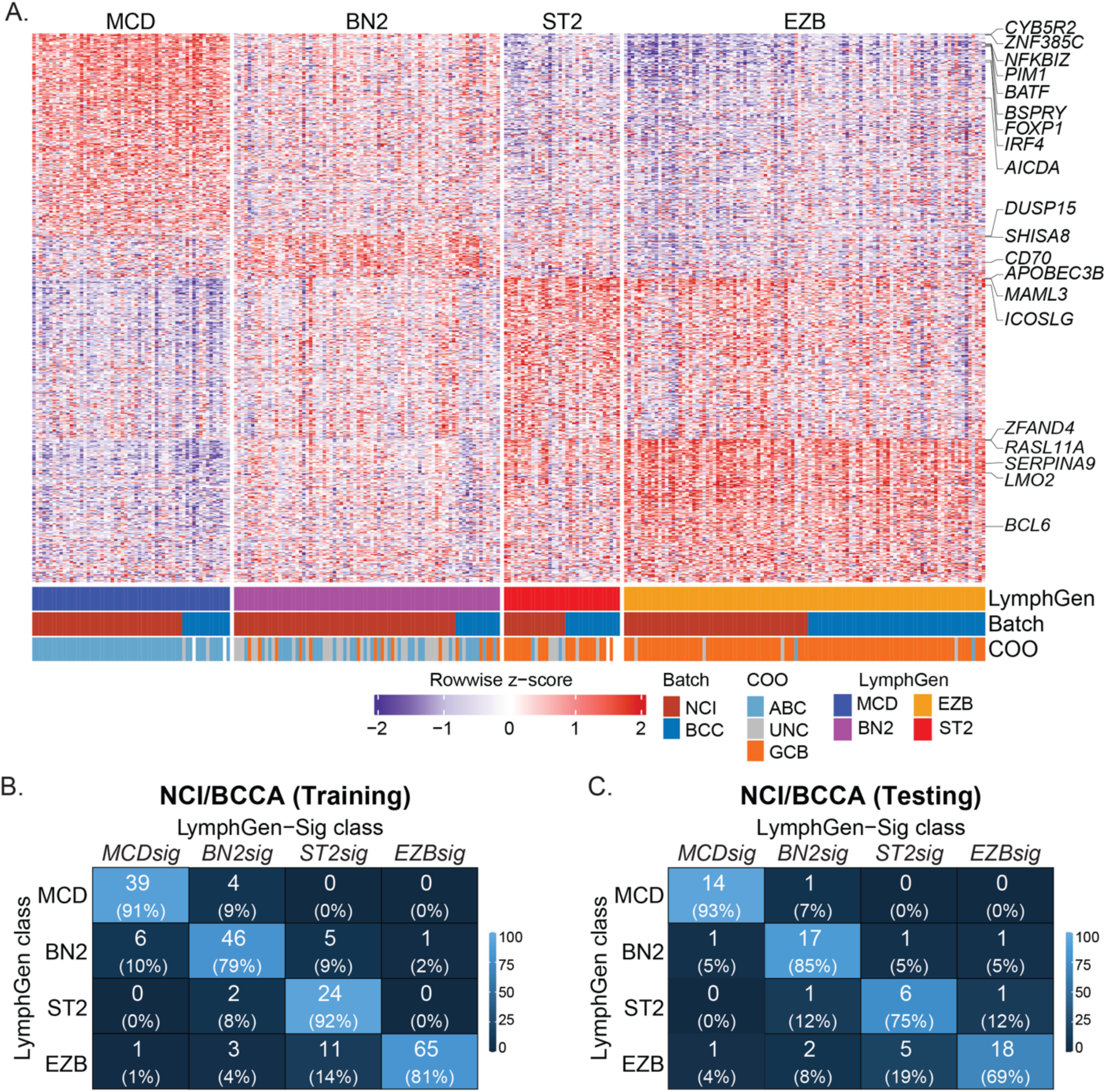
LG subtypes are accurately recapitulated by transcriptional features. **A.** Heatmap of subtype-specific gene expression signatures across DLBCL cases in the NCI/BCCA cohort. Expression values are shown as row-wise *z*-scores. Samples are annotated by LymphGen (LG) class, batch (NCI or BCCA), and cell of origin (COO). Selected representative genes are indicated. **B–C.** Confusion matrices showing concordance between LymphGen genetic classes and transcriptional LGsig classes in the NCI/BCCA training **(B)** and testing **(C)** cohorts. Numbers indicate case counts, with percentages shown in parentheses.

Using these features, we trained a nearest shrunken centroid classifier on the NCI/BCCA dataset with a 75:25 training-testing split to determine the extent to which LG subtypes could be accurately classified by transcriptional features alone. To refine the model, we (1) performed 5-fold cross validation to determine the optimal shrinkage and (2) systematically varied the maximum number of DGE features included, ranging from 250 to 600. Because some DGE comparisons contain overlapping genes and some comparisons may yield fewer significant genes than the maximum, the effective number of genes retained in the final model was smaller than the specified maximum at each threshold. Model performance, assessed by accuracy, balanced accuracy, macro-F1, and weighted-F1, was broadly consistent across this range, with the 300-gene maximum threshold model showing the highest average metrics on the test set (accuracy 0.80, balanced accuracy 0.81, macro-F1 0.78, weighted-F1 0.80) **(Supplementary Figure 2A)**. However, given the potential for overfitting with smaller feature sets, we selected the model initialized with a 450-gene maximum threshold (294 final genes) as our classifier (**Supplementary Table 1**). This model maintained strong performance (accuracy 0.80, balanced accuracy 0.80, macro-F1 0.78, weighted-F1 0.80) while providing greater robustness and stability across cross validation folds **(Supplementary Figure 2B)**.

The final classifier – termed LymphGen-sig (LGsig) - demonstrated strong subtype-specific performance **(Figure 1B–C)**. Moving forward, we refer to genetically defined LG subtypes as *bona fide (bf)*-MCD, *bf*-BN2, *bf*-ST2, and *bf*-EZB, and to transcriptionally-defined LGsig assignments as *MCDsig*, *BN2sig*, *ST2sig*, and *EZBsig*. Each LGsig subtype represents a transcriptional category that includes corresponding *bona fide* LG cases together with reassigned genetically unclassified LG cases (e.g. u-*MCDsig*) and cross-classified cases (e.g x-*MCDsig*, which are cases genetically assigned to one LG subtype but transcriptionally reassigned by LGsig to another, **Supplementary Figure 2C**). In the training set, 91% of *bf*-MCD cases were classified as *MCDsig*, 79% of *bf*-BN2 as *BN2sig*, 92% of *bf*-ST2 as *ST2sig*, and 81% of *bf*-EZB as *EZBsig* **(Figure 1B)**. Similar performance was observed in the test set **(Figure 1C)**. Cross-classifications were most frequent for EZB into *ST2sig*, while MCD DLBCLs were consistently identified with the highest accuracy into *MCDsig*. Taken together, these results show that LG subtypes are transcriptionally distinct, and that LGsig reliably classifies genetically defined LG cases using gene expression data alone.

### LG-unclassified DLBCLs are assigned into LGsig subtypes based on shared transcriptional features

The inability to classify ∼40% of DLBCLs is a major limitation of conventional LG.^11^ We hypothesized that LG-unclassified DLBCLs may nonetheless engage similar downstream transcriptional programs as genetically defined (*bf-*LG) subtypes, but lack the genetic alterations required to support a high probability LG assignment. Application of LGsig to the LG-unclassified and -composite groups within the NCI/BCCA dataset demonstrated that these cases were robustly transcriptionally aligned with specific *bf*-LG subtypes (**Figure 2A-B)**. Whereas LG leaves ∼40% of DLBCLs unclassified in the NCI/BCCA dataset, LGsig assigned every case, with LG-unclassified and LG-composite tumors distributed across *MCDsig*, *BN2sig*, *ST2sig*, and *EZBsig* classes. Among LG-unclassified DLBCLs, 9% were classified as *EZBsig*, 37% as *BN2sig*, 30% as *MCDsig*, and 23% as *ST2sig*. Similarly, LG-composite cases were redistributed across the four subtypes, with 16% assigned as *MCDsig* and 32% *BN2sig*, 34% *EZBsig*, and 18% *ST2sig*.

**Figure 2.**
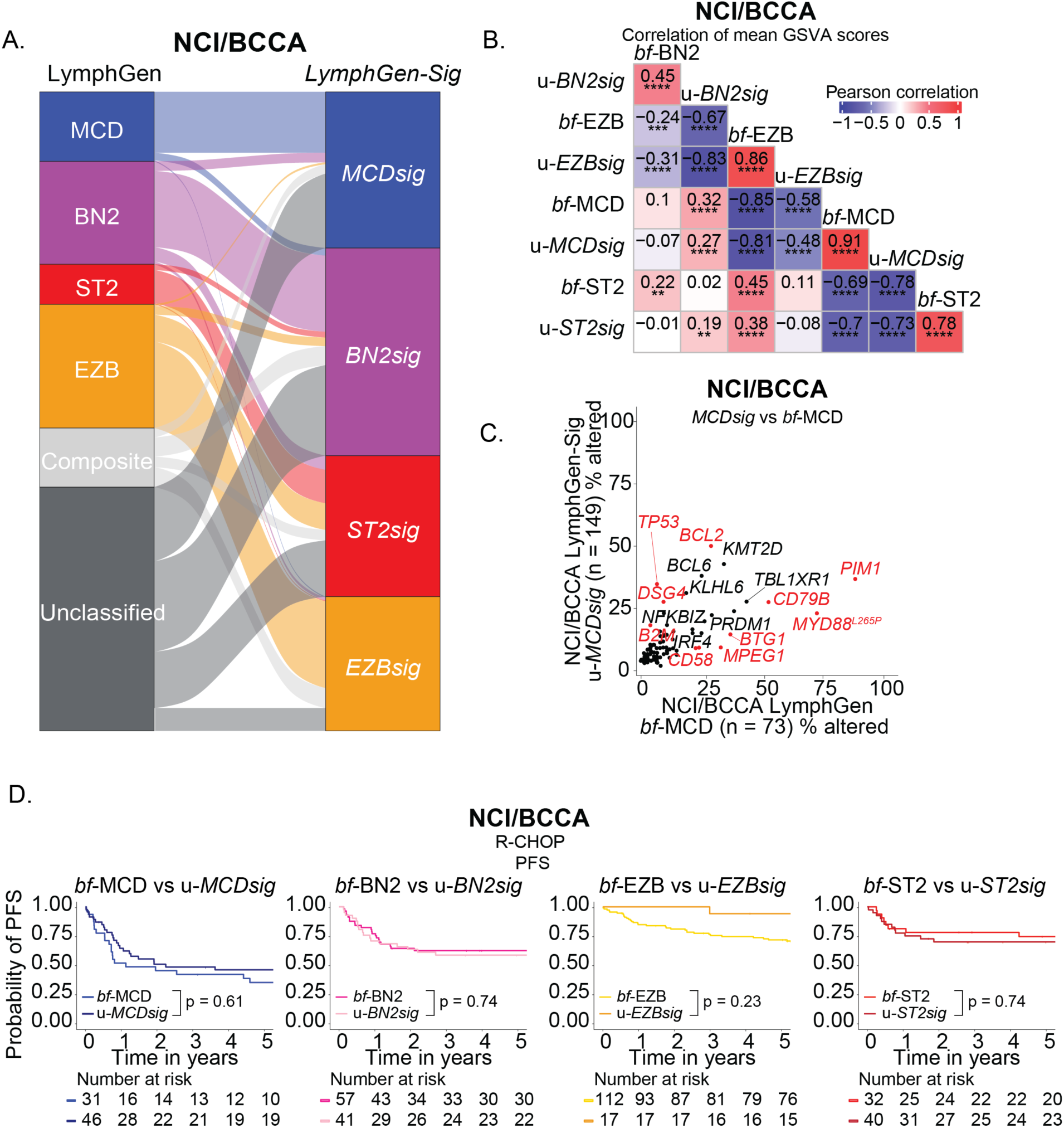
LG-unclassified DLBCLs are assigned into LGsig subtypes based on shared transcriptional features. **A.** Alluvial plot showing the relationship between LymphGen genetic classes and LGsig transcriptional classes in the NCI/BCCA cohort. **B.** Correlation matrix showing pairwise correlations of mean GSVA scores between LymphGen genetic classes and LGsig transcriptional classes (excluding bf-LG cases) in the NCI/BCCA cohort. P-values were computed using a Pearson correlation t-test and adjusted for multiple testing using the Benjamini-Hochberg procedure. Adjusted p-values are shown (* adj p < 0.05, ** adj p < 0.01, ***adj p < 0.001, **** adj p < 0.0001). **C.** Scatter plot comparing the proportion of cases with genomic alterations in individual genes in *MCDsig* DLBCLs (y-axis, excluding bf-MCD DLBCLs) compared with *bf*-MCD DLBCLs (x-axis) in the NCI/BCCA cohort. Dots highlighted in red are significantly enriched (Fisher’s exact test adj p < 0.1). **D.** Kaplan–Meier curves showing progression-free survival (PFS) for R-CHOP–treated patients stratified by LymphGen genetic class and corresponding LGsig transcriptional class (excluding bf-LG classifiable DLBCLs) in the NCI/BCCA cohort. P-values indicate comparisons between each LGsig group and its corresponding genetic subtype.

Although LGsig subtypes were defined using 294 selected features, we evaluated whether u-LGsig cases resembled their *bona fide* genetically-defined counterparts at a whole-transcriptome level. To systematically compare transcriptome-wide similarity, we correlated mean gene set variation analysis (GSVA) scores for 223 lymphoma-related genesets across *bf*-LG and u-LGsig subtypes **(Figure 2B)**. For each subtype, the strongest correlations were observed between *bf*-LG and u-LGsig cases of the same class (e.g., *bf*-MCD vs u-*MCDsig*, *bf*-EZB vs u-*EZBsig*). Correlations were highest for *bf-*MCD and u-*MCDsig* (r = 0.91, p = 0.0076), *bf-*EZB and u-*EZBsig* (r = 0.86, p = 0.015), and *bf-*ST2 and u-*ST2sig* cases (r = 0.78, p = 0.014), while *bf-*BN2 and u-BN2sig DLBCLs also demonstrated positive, but more modest, correlation (r = 0.45, p = 0.21). By contrast, correlations between mismatched subtypes were consistently weaker, supporting the specificity of LGsig assignments **(Figure 2B, Supplementary Figure 3A-D)**.

Having established that LGsig faithfully identifies LG-unclassified DLBCLs that recapitulate the transcriptional programs of genetically defined LG subtypes, we next evaluated whether LG-unclassified tumors also exhibited the characteristic genetic features of their corresponding *bf*-LymphGen subtype. To do so, we compared the mutational profiles of *bf*- and the corresponding u-LGsig DLBCLs **(Figure 2C and Supplementary Figure 4A-C)**. As expected, *bf*-LymphGen subtypes were enriched for their hallmark lesions, consistent with their genetic definitions, whereas these canonical drivers were markedly less frequent in u-LGsig tumors. For example, whereas 75% of *bf*-MCDs carried *MYD88*^L265P^ mutations and 55% harbored *CD79B* alterations, only 23% and 28% of u-*MCDsig* cases carried these alterations, respectively, despite sharing similar transcriptional programs **(Figure 2C)**. Similarly, *BCL6* alterations and *NOTCH2* alterations were present in 75% and 31% of *bf*-BN2 cases but only 42% and 3% of u-BN2sig tumors, respectively **(Supplementary Figure 4A)**. Among *bf*-EZB tumors, 47% carried *EZH2* alterations and 75% harbored *BCL2* alterations, compared to much lower frequencies in u-*EZBsig* tumors (28% and 56% respectively) **(Supplementary Figure 4B)**. Finally, in the *ST2sig* class, *bf-*ST2 DLBCLs showed higher rates of *SGK1* and *TET2* alterations relative to their u-ST2sig counterparts (*SGK1:* 52% vs 24%; *TET2:* 24% and 7%; **Supplementary Figure 4C).** Together, these results demonstrate that LGsig augments conventional LG classification by identifying additional DLBCLs that lack canonical driver mutations yet converge on similar downstream transcriptional states that contribute to subtype identity.

Lastly, we asked whether transcriptionally defined LGsig subtypes recapitulated the prognostic associations of LG classifications in the setting of front-line R-CHOP chemotherapy. U-LGsig DLBCLs exhibited PFS and OS patterns nearly identical to their *bf*-LymphGen counterparts **(Figure 2D and Supplementary Figure 4D)**. Thus, LGsig identifies LymphGen-unclassified DLBCLs that broadly share transcriptional and clinical features with *bf*-LymphGen subtypes, which supports their reclassification using the LGsig algorithm.

### LGsig redefines the A53 LG subtype

Although the A53 subtype has been considered a distinct entity within the LG framework – defined largely by chromosomal copy number alterations and *TP53* mutations – transcriptional analyses revealed substantial heterogeneity when compared to other LG subtypes. Using LGsig, we found that 60% of A53 tumors aligned with the *MCDsig* group in the NCI/BCCA dataset, while the remainder distributed across *BN2sig*, *ST2sig*, and *EZBsig* **(Figure 3A-B)**. Cross-classified A53 (x-A53) DLBCLs showed transcriptional patterns highly similar to their corresponding *bf*-LG subtype **(Figure 3C and Supplementary Figure 5A-D)**. As the majority of A53-DLBCLs were reclassified as *MCDsig*, we focused our subsequent analyses on this subset. Interestingly, although A53-*MCDsig* cases were nearly indistinguishable from *bf*-MCDs at a transcriptome-wide level (r = 0.89, p = 0.004; **Figure 3C**), their genetic profiles diverged. Hallmark *MYD88/CD79B* mutations were less frequent than in *bf*-MCD DLBCLs, and *A53-MCDsig* tumors exhibited selective enrichment for other alterations including *TP53* and *BCL2* lesions **(Figure 3D)**. Additionally, mutations in genes not classically associated with the MCD subtype, such as *NFKBIZ,* were enriched in the A53-*MCDsig* DLBCLs, suggesting they may represent alternative drivers of the MCD transcriptional program in A53 DLBCLs. *NFKBIZ* encodes the inducible nuclear protein IκBζ, a transcriptional regulator that amplifies NF-κB–dependent programs and has been implicated in ABC-DLBCL biology. Thus, its enrichment in MCD-like A53 tumors is consistent with convergence on the NF-κB–driven program that defines the MCD subtype.

**Figure 3.**
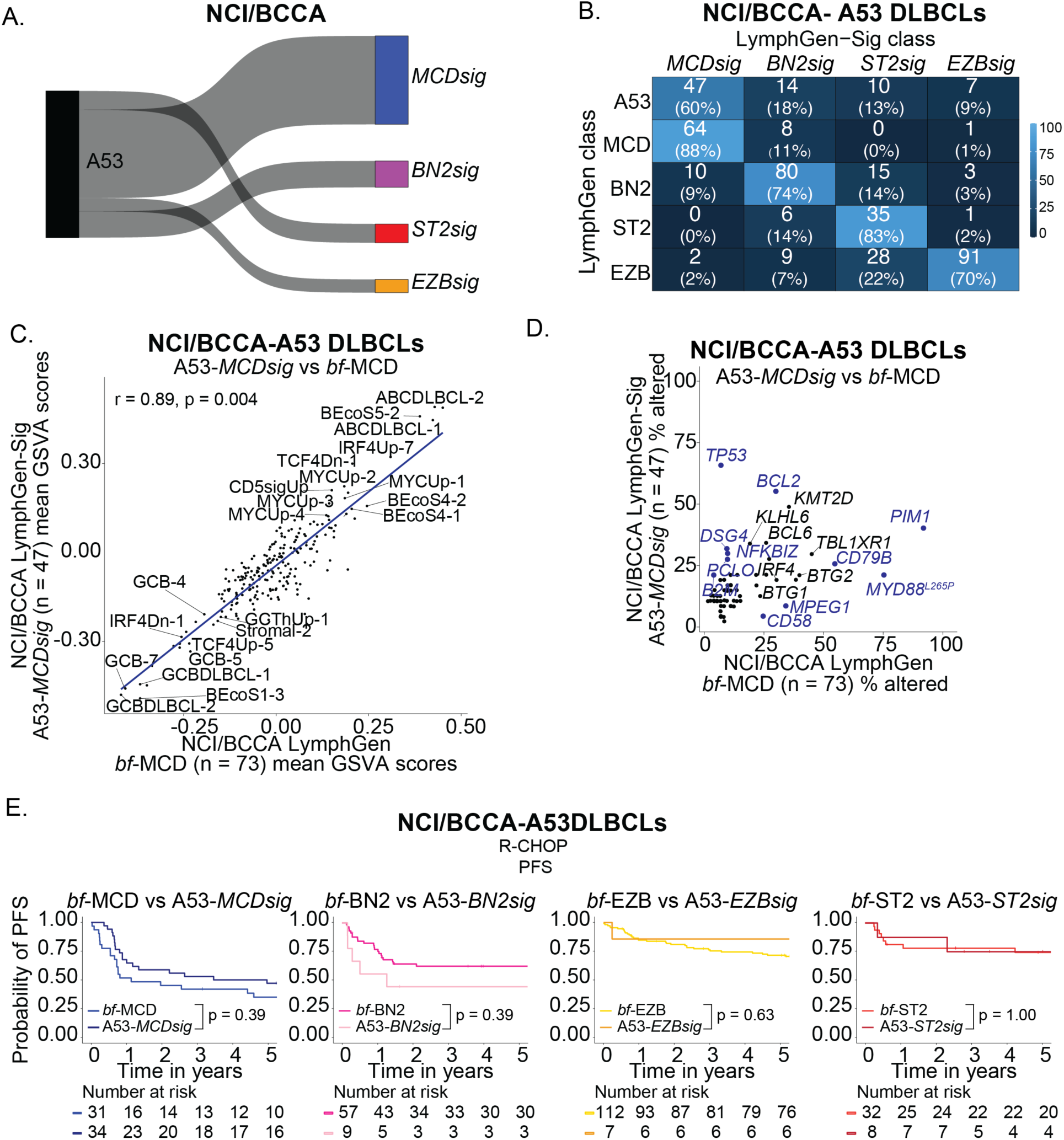
LGsig reassigns A53 LG DLBCLs. **A.** Alluvial plot showing the relationship between A53 DLBCLs and LGsig transcriptional classes in the NCI/BCCA cohort. **B.** Confusion matrices showing concordance between LymphGen genetic classes and transcriptional LGsig classes in the NCI/BCCA training, with A53 DLBCLs included. **C.** Scatter plots showing the mean GSVA scores for selected lymphoma-related gene sets for *bf-*MCD DLBCLs (x-axis) and A53 DLBCLs assigned to the *MCDsig* cluster in the NCI/BCCA dataset (y-axis). **D.** Scatter plot comparing the proportion of cases with genomic alterations in individual genes in A53- *MCDsig* DLBCLs (y-axis) compared with *bf*-MCD DLBCLs (x-axis) in the NCI/BCCA cohort. Dots highlighted in blue are significantly enriched (Fisher’s exact test adj p < 0.1). **E.** Kaplan–Meier curves showing progression-free survival (PFS) for R-CHOP–treated patients stratified by LymphGen genetic class and A53-LGsig subtype in the NCI/BCCA cohort. P-values indicate comparisons between each A53-LGsig group and its corresponding LG genetic subtype.

Clinically, the outcomes of reclassified A53 DLBCLs treated with R-CHOP mirrored the outcomes of their corresponding *bf-*LG counterparts. For example, the PFS and OS of A53-*MCDsig* patients treated with R-CHOP mirrored *bf*-MCD DLBCLs and trended towards inferior outcomes compared to other A53 tumors **(Figure 3E and Supplementary Figure 5E)**. These findings suggest that A53 is not a uniform DLBCL subtype, but rather a biologically heterogeneous collection of cases that can be classified into established transcriptional subtypes using LGsig. In this sense, these data indicate LGsig complements genetic classification by uncovering underlying transcriptional identity that may be obscured when considering genetic alterations alone.

### Comparison of LGsig to other molecular DLBCL classifiers

To contextualize LGsig among other transcriptional frameworks, we compared it with established classifiers, including COO,^8^ DZsig,^14,16^ and Ecotyper B-cell states,^18^ as well as CD5sig,^22^ a signature we recently defined for CD5⁺ DLBCL. *MCDsig* was enriched within the activated B-cell (ABC) COO (90%) and captured 95% of CD5sig⁺ tumors, while *EZBsig* aligned with the germinal center B-cell (GCB) COO (95%) and strongly overlapped with DZsig⁺ cases **(Supplementary Figure 6A-B)**. LGsig provided additional resolution of COO by partitioning ABC cases into two distinct groups (*MCDsig* and *BN2sig*) and dividing GCB DLBCLs into EZBsig and ST2sig. COO-unclassified tumors were mainly distributed across *BN2sig* and *ST2sig* LGsig subtypes **(Supplementary Figure 6A)**. Although CD5sig⁺ tumors were distributed almost entirely within *MCDsig*, the latter also included many CD5sig⁻ cases, underscoring that LGsig captures a broader transcriptional program **(Supplementary Figure 6B)**. Similarly, *BN2sig* showed no strong overlap with any single classifier **(Supplementary Figure 6A-B)**. Finally, Ecotyper B-cell state analysis revealed preferences for certain LGsig classes (*EZBsig* enriched for S1, *MCDsig* for S5) but also heterogeneity within subtypes **(Supplementary Figure 6C)**. These findings indicate that LGsig is partially related to existing transcriptional classifiers yet distinct in its ability to unify and extend them, capturing subtype-level programs that integrate across multiple biological features rather than being limited to a single signature or dimension of classification.

Comparison with DLB*class*, a recently developed genetic classifier that assigns all DLBCLs to one of five genetic classes (C1-C5), revealed partial overlap between DLB*class* and LGsig classifications **(Supplementary Figure 6D)**.^19^ C1 and C5 tumors predominantly mapped to *BN2sig* and *MCDsig*, respectively, while C3 and C4 were enriched for *EZBsig* and *ST2sig*. However, C2 - characterized by large-scale copy number alterations and *TP53* mutations - were equally distributed across all four LGsig subtypes similar to our findings with the A53 LG subtype that is analogous to the C2 DLB*class* **(Supplementary Figure 6D and Figure 3A)**. These data indicate relatively strong overlap between DLB*class* and LGsig, while revealing that C2 DLBCLs comprise a transcriptionally heterogenous group with distinct subtype-specific transcriptional programs that can be readily uncovered by LGsig. Given the lack of therapeutic strategies to target *TP53* alterations, this added resolution may help identify alternative, targetable pathways relevant for precision medicine strategies in A53/C2 DLBCLs.

### LGsig identifies LG-unclassified DLBCLs that benefit from pola-based chemoimmunotherapy in POLARIX

In order to determine its clinical relevance, we applied LGsig to archival DLBCL tumors from the POLARIX study, in which patients with previously untreated DLBCL (IPI 2-5) were randomized to receive treatment with pola-R-CHP or R-CHOP.^3,23^ The distribution of LG and LGsig classifications in the POLARIX cohort is shown in **Supplementary Figure 7A**. Overall, LGsig exhibited similar performance in accurately classifying *bf*-LG DLBCLs as observed in our training and testing datasets, with 78.5% balanced accuracy **(Figure 4A)**. In POLARIX, 57.1% of DLBCLs were classified by LG versus 100% by LGsig **(Figure 4B)**. Similar to our discovery cohort, u-LGsig DLBCLs demonstrated broad transcriptional similarity to *bf*-LG DLBCLs assigned to the same class (i.e. u-*MCDsig* compared to *bf*-MCD) **(Supplementary Figure 7B-F)**. Together, these results validate the performance of the LGsig model.

**Figure 4.**
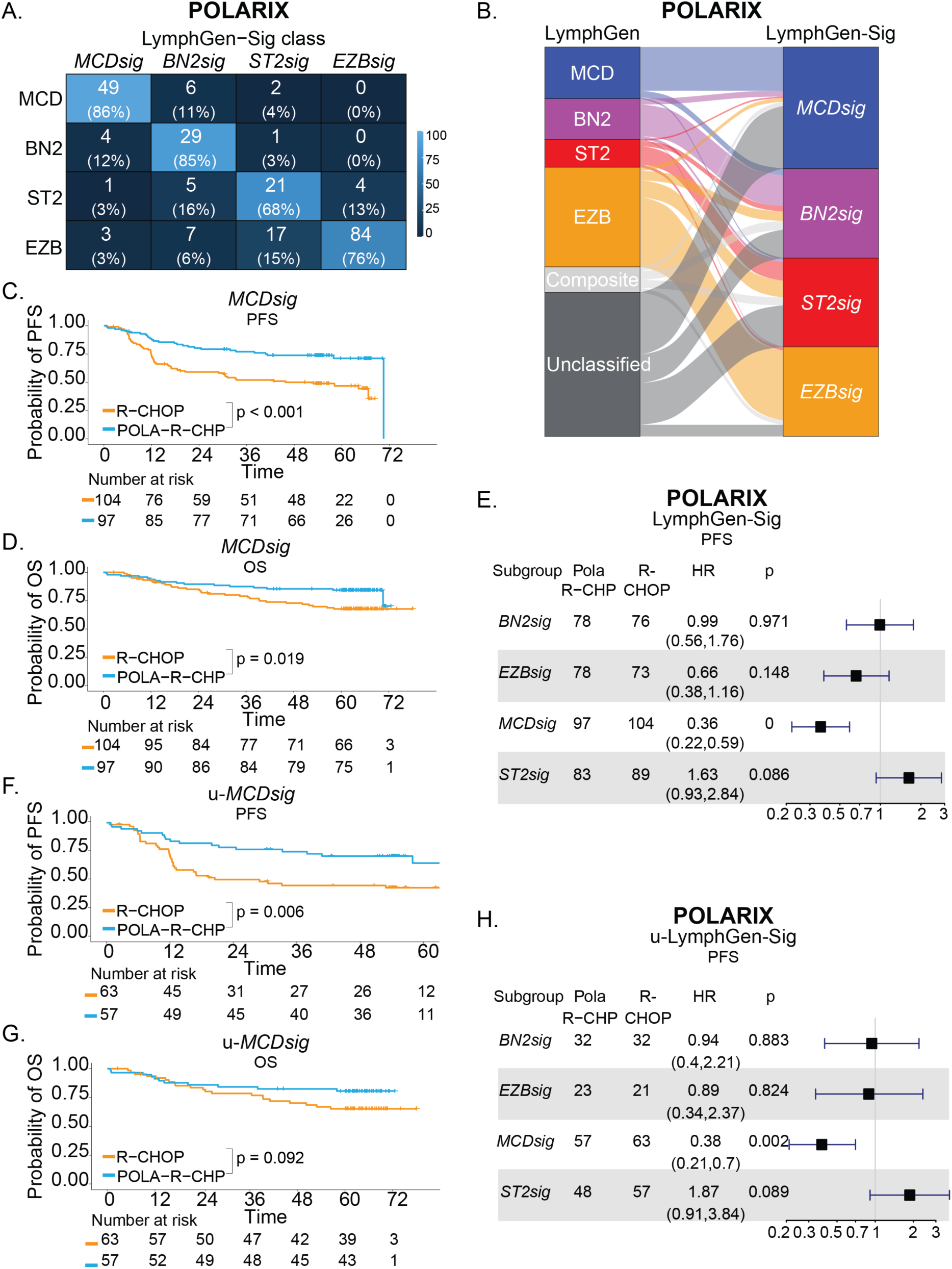
LGsig identifies LG-unclassified DLBCLs that benefit from pola-based chemoimmunotherapy in POLARIX. **A.** Confusion matrix showing concordance between LymphGen genetic classes and LGsig transcriptional classes in the POLARIX cohort. Values indicate case counts with row percentages in parentheses. **B.** Alluvial plot showing the relationship between LymphGen genetic classes and LGsig transcriptional classes in the POLARIX cohort. **C–D.** Kaplan–Meier curves comparing progression-free survival (C) and overall survival (D) between R-CHOP- and pola–R–CHP-treated *MCDsig* DLBCLs cases in the POLARIX cohort. Numbers at risk are shown below each plot. Unadjusted p-values are displayed. **E.** Forest plot showing progression-free survival hazard ratios for pola–R–CHP versus R-CHOP-treated DLBCLs across LGsig subgroups in the POLARIX cohort. Hazard ratios are adjusted for International Prognostic Index score (2 vs 3–5), age (<60 vs ≥60 years), and COO (activated B cell, germinal center B cell, unclassified, unknown). **F–G.** Kaplan–Meier curves comparing progression-free survival (F) and overall survival (G) between R-CHOP and pola–R–CHP-treated DLBCLs in u-*MCDsig* cases in the POLARIX cohort. Numbers at risk are shown below each plot. Unadjusted p-values are displayed. **H.** Forest plot showing progression-free survival hazard ratios for pola–R–CHP versus R-CHOP-treated DLBCLs across u-LGsig subgroups in the POLARIX cohort. Hazard ratios are adjusted for International Prognostic Index score (2 vs 3–5), age (<60 vs ≥60 years), and COO (activated B cell, germinal center B cell, unclassified, unknown).

Regarding clinical outcomes, the PFS and OS for LG subtypes in the R-CHOP and pola-R-CHP treatment arms of POLARIX are shown in **Supplementary Figure 8A-H,** while the corresponding analysis for LGsig-defined subtypes are shown in **Figure 4C-D and Supplementary Figure 9A-F**. Among LG subtypes, only MCD DLBCLs demonstrated a significant PFS benefit from pola-R-CHP over R-CHOP (HR 0.32, 95% CI: 0.12-0.81; P=0.016; **Supplementary Figure 8A-B**). Of LGsig subtypes, *MCDsig* DLBCLs exhibited improved PFS and OS with pola-R-CHP compared to R-CHOP, a finding that retained statistical significance after adjusting for age, international prognostic index score, and COO (PFS: HR 0.36, 95% CI: 0.22-0.59; P < 0.001 and OS: HR 0.43, 95% CI: 0.22-0.81; P=0.009; **Figure 4C-E**). Other LGsig subtypes (BN2sig, EZBsig, ST2sig) demonstrated non-significant PFS results (**Figure 4E, Supplementary Figures 9A-G)**. Lastly, although LG-unclassified DLBCLs did not benefit from pola-R-CHP compared to R-CHOP, those LG-unclassified cases that were specifically reassigned to the *MCDsig* cluster had improved PFS (HR 0.38, 95% CI: 0.21-0.7; P=0.002) and OS (HR 0.48, 95% CI: 0.23-1.01; P=0.055) with pola-R-CHP over R-CHOP after adjusting for age, international prognostic index score, and COO similar to *bf*-MCD DLBCLs **(Figure 4F-H and Supplementary Figure 10A-G)**. Thus, these data indicate that relying on genetic classification alone may not fully capture biologically related subsets of DLBCLs that similarly benefit from relevant front-line therapies, and that LGsig extends genetic classification by identifying previously unclassified DLBCLs with subtype-specific transcriptional programs that predict sensitivity to pola-based chemo-immunotherapy.

## Discussion

Here, we demonstrate that genetic DLBCL subtypes defined by the LG framework can be accurately identified using transcriptional features alone. The LGsig classifier, trained on a 294-gene feature set, identified *bona fide* MCD, BN2, ST2, and EZB subtypes with high accuracy and extended classification to ∼40% of DLBCLs categorized as unclassified or composite by LG. These cases reassigned by LGsig mirrored transcriptome wide gene expression programs and clinical outcomes of their genetically defined counterparts, indicating that transcriptional states can stratify DLBCL into biologically and therapeutically relevant classes in a manner not captured using genetic data alone. LGsig enhances DLBCL genetic classification by increasing the proportion of samples that can be meaningfully classified, while refining LG categories such as A53, where transcriptional reclassification resolves biological and clinical heterogeneity.

These findings position LGsig as a next-generation DLBCL classifier that builds upon and integrates insights from existing genetic classifiers. LG established a framework for clustering DLBCLs based on co-occurring genomic alterations, which has ultimately resolved heterogeneity beyond COO, and offered an improved approach for delivering precision medicine in this disease.^11,20^ However, LG leaves approximately 40% of DLBCLs unclassified, which is problematic for its potential use in the clinic. Rather than utilizing genetic data, LGsig leverages bulk RNA sequencing data alone to define transcriptional states that reflect underlying genetic programs, which may facilitate its clinical translation. Moreover, like LG, it resolves the heterogeneity of COO by subclassifying ABC DLBCLs into MCD- and BN2-like groups, and GCB DLBCLs into EZB- and ST2-like groups. However, unlike LG, it defines these relationships through transcriptional states. This importantly permits the identification of non-genetic pathways that contribute to shared DLBCL biology, which facilitates classification of all cases. Thus, LGsig provides a unified framework that leverages insights from both genetic and non-genetic components of disease biology, thereby advancing toward a more robust molecular taxonomy of DLBCL.

Extending this principle, our findings highlight that DLBCL subtypes can be transcriptionally defined even when canonical genetic drivers are absent. For instance, many LG-unclassified and LG-A53 DLBCLs lacked hallmark alterations such as *MYD88* or *CD79B* yet still converged on the same MCD transcriptional program. This convergence may reflect 1) alternative, non-canonical mutations, 2) non-coding mutations better captured by whole genome sequencing, or 3) non-genetic mechanisms. As an example of the former, A53 DLBCLs that were cross-classified as *MCDsig* showed enrichment for *NFKBIZ* amplifications. *NFKBIZ* encodes IκBζ, a nuclear co-activator that promotes NF-κB–dependent transcription and contributing to the MCD-like transcriptional program in these DLBCLs.^25,33,34^ In addition to copy number gains, *NFKBIZ* can be dysregulated through noncoding 3′ UTR mutations that stabilize its transcript and enhance IκBζ expression.^25^ Since LymphGen is based on whole exome and not whole genome sequencing, such noncoding variants may not be detected, suggesting that their contribution to NF-κB activation—and thus to the MCD-like transcriptional state—may be underappreciated by current genomic classifiers. Beyond non-canonical LG genetic alterations, sustained BCR signaling itself can also represent a powerful non-genetic mechanism driving similar NF-κB transcriptional activation. Chronic BCR activation can arise from structural modifications of the receptor, altered glycosylation patterns, or microenvironmental engagement, leading to persistent NF-κB pathway activity even in the absence of canonical MCD pathway mutations.^35–38^ Such processes highlight that transcriptional states identified by LGsig may reflect functional signaling convergence rather than strictly genetic causation, enabling identification of biologically coherent subtypes that lie beyond the reach of mutation-based classifiers. In support of this idea, recent multimodal single-cell profiling demonstrated reproducible subtype-associated gene expression programs in malignant B cells across LymphGen subtypes and further showed that LG-unclassified DLBCLs can transcriptionally align with these programs.^39^ Notably, this analysis also revealed substantial transcriptional overlap between A53 and MCD DLBCLs, consistent with our finding that A53 cases can be transcriptionally reclassified and mirror the biology of the groups to which they align. Together, these data underscore the value of LGsig in uncovering distinct therapeutic vulnerabilities within LG-unclassified and A53 DLBCLs that are not captured by genetic classifiers, such as supporting the application of pola-based therapy for those adopting a MCD signature given the clear survival benefit with pola-R-CHP over R-CHOP for *MCDsig* DLBCLs in the POLARIX study.

This study has limitations. LGsig was trained on bulk RNA-seq data, and its performance on alternative platforms, including microarray or targeted expression assays, remains to be determined. Cohort size and composition also posed constraints, as rare subtypes such as N1 were excluded due to limited representation, and larger datasets will be essential to refine the classifier. Accuracy of predicted LG assignments also varied by subtype, which could also improve with application of the platform to larger datasets and model training. The current model does not incorporate a probability cutoff, which may limit flexibility in handling borderline assignments. In addition, while LGsig integrates with existing transcriptional and genetic frameworks, it has not yet been systematically benchmarked against all available classification schemes, nor incorporated with immune classifiers that may provide complementary prognostic information in the setting of immunotherapies such as bispecific antibodies and CAR T cells.^40,41^ Finally, while we demonstrate the utility of LGsig as a biomarker for pola-based therapy, further studies are warranted to investigate the impact of LGsig subtype classifications on outcomes to other targeted therapies such as BTK inhibitors and BCL6 degraders, as these therapies each target oncogenic pathways specifically activated within certain LGsig subtypes.

Despite these caveats, LGsig represents a significant advance. It complements genetic classification by expanding subtype assignment to include previously unclassified DLBCLs, reassigning A53 into biologically and clinically relevant categories, and uncovering a subset of LymphGen-unclassified cases that benefit from pola-R-CHP in the POLARIX study. Furthermore, LGsig can prospectively classify DLBCL cases, supporting its translational potential as a platform to guide therapeutic decisions in future precision medicine trials. Taken together, these findings demonstrate that transcriptional classifiers can extend the reach of genetic frameworks and provide actionable insights for precision medicine in DLBCL.

## Data Sharing

Requests for access to the data from the POLARIX study can be directed to Roche. For eligible studies, qualified researchers may request access to individual patient-level clinical data for POLARIX through a data request platform. At the time of writing, this request platform is Vivli (https://vivli.org/ourmember/roche/). For-up-to-date details on Roche’s Global Policy on the Sharing of Clinical Information and how to request access to related clinical study documents, see here: https://go.roche.com/data_sharing. Anonymized records for individual patients across more than one data source external to Roche cannot, and should not, be linked because of a potential increase in risk of patient reidentification. Genomic data from BCCA and NCI cohorts can be requested as outlined within the manuscript of each of these respective studies.

## Acknowledgements

The authors sincerely thank the patients and their families for their invaluable participation in the POLARIX study and gratefully acknowledge Genentech for their collaboration in providing genomic data from archived tumor samples collected in POLARIX. This work was supported by the American Society of Hematology Scholar Award (J.K.G.).

## Author Contributions

**Conception and design:** Alan Cooper, Sravya Tumuluru, Justin Kline, James K. Godfrey

**Financial support:** Justin Kline, James K. Godfrey

**Administrative support:** Justin Kline, James K. Godfrey

**Provision of study materials or patients:** Yanwen Jiang, Connie Lee Batlevi, Will Harris, Gilles Salles, Marek Trneny, Georg Lenz, Franck Morschhauser, Fabrice Jardin, Alex F. Herrera, Justin Kline, James K. Godfrey

**Collection and assembly of data:** Alan Cooper, Sravya Tumuluru, Yanwen Jiang, Connie Batlevi, Will Harris, Sandhya Balasubramanian, Matthew Sugidono, Alex F. Herrera, Justin Kline, James K. Godfrey

**Data analysis and interpretation:** Alan Cooper, Sravya Tumuluru, Yanwen Jiang, Connie Batlevi, Will Harris, Alex F. Herrera, Justin Kline, James K. Godfrey

**Manuscript writing:** All authors

**Final approval of manuscript:** All authors

**Accountable for all aspects of the work:** All authors

---

**Supplementary Figure 1.**
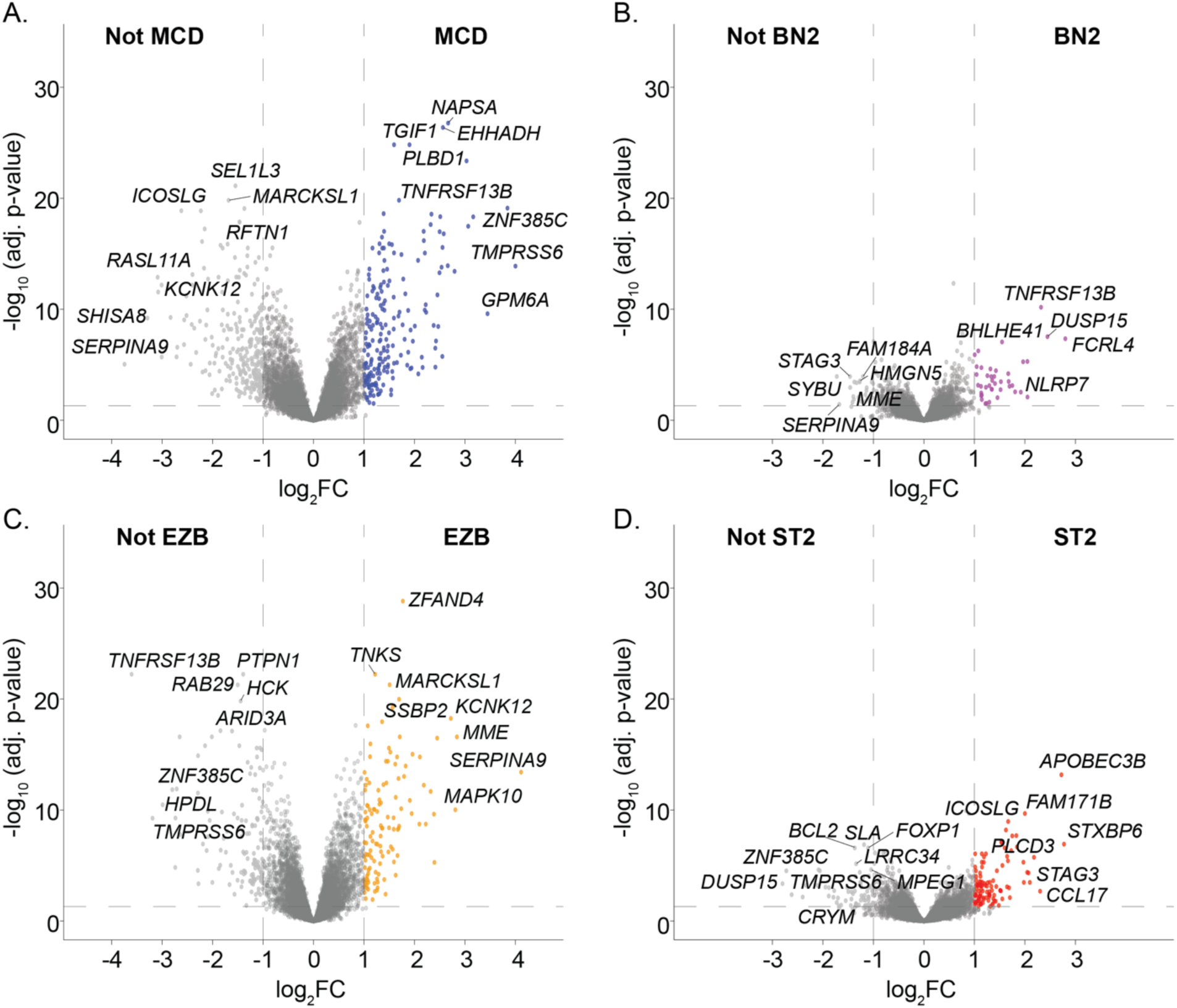
LG subtypes are accurately recapitulated by transcriptional features. **A–D.** Volcano plots of differentially expressed genes (DEGs) comparing each *bf*-LymphGen subtype with all other LG-classifiable DLBCL cases in the NCI/BCCA cohort. **A.** MCD vs all other subtypes **B.** BN2 vs all other subtypes **C.** EZB vs all other subtypes **D.** ST2 vs all other subtypes. The x-axis shows log₂ fold change (vertical dashed lines indicate log₂FC = ±1, horizontal dashed line indicates adj p-value < 0.05) and the y-axis shows −log₁₀ adjusted p-value. Selected significantly up- or down-regulated genes are labeled.

**Supplementary Figure 2.**
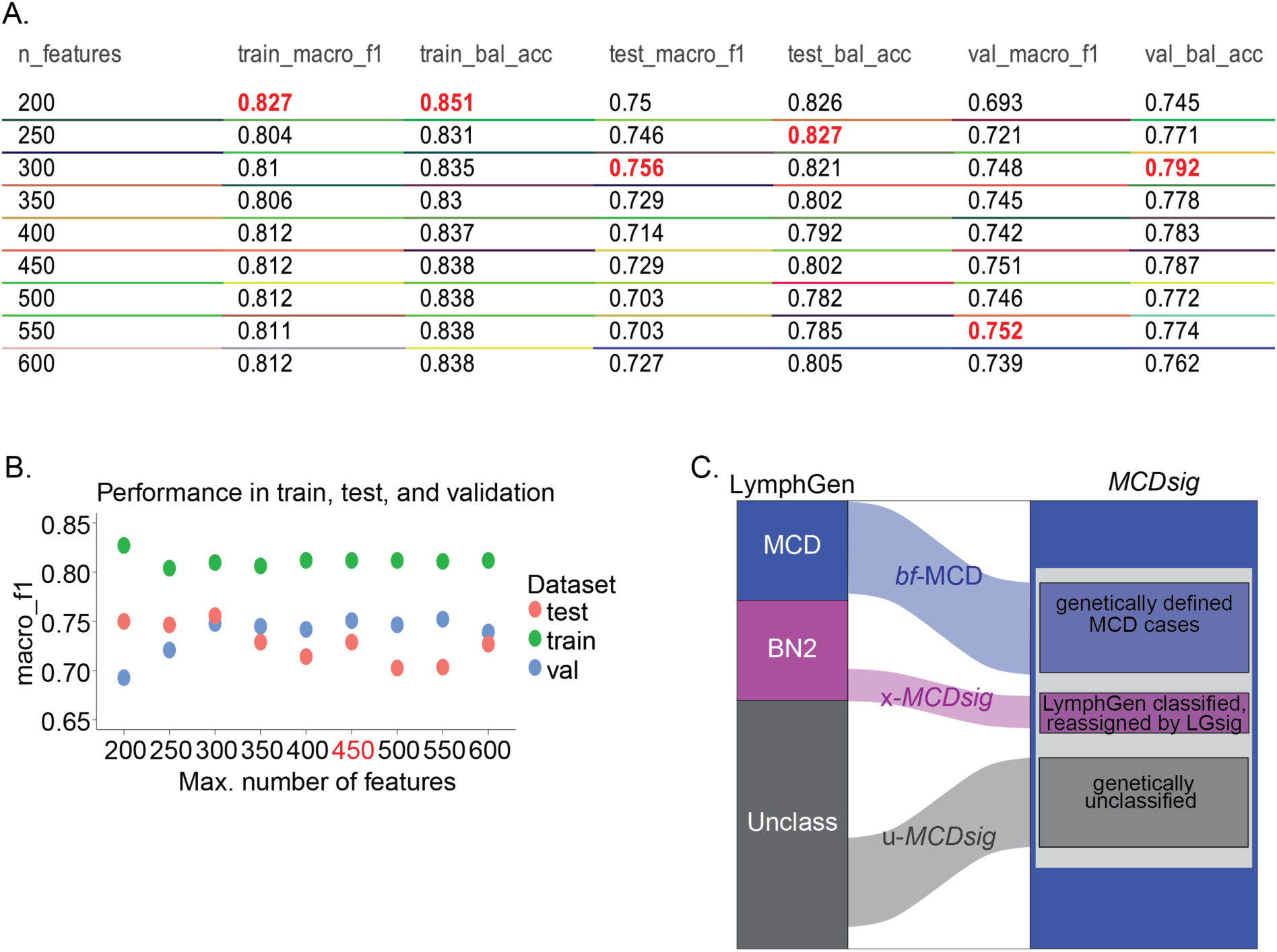
LG subtypes are accurately recapitulated by transcriptional features. **A.** Performance metrics for nearest shrunken centroid classifiers trained with varying maximum numbers of gene features (200–600). Shown are macro-F1 score and balanced accuracy for training, test, and validation datasets. Values highlighted in red indicate the best-performing model for each metric. **B.** Macro-F1 scores across training (green), test (red), and validation (blue) datasets as a function of the maximum number of features supplied to the classifier. The 450 maximum threshold model (highlighted in red) was selected for downstream analyses. **C.** Representative schematic illustrating the relationship between genetically defined LymphGen subtypes and transcriptionally defined LGsig categories, shown using *MCDsig* as an example.

**Supplementary Figure 3.**
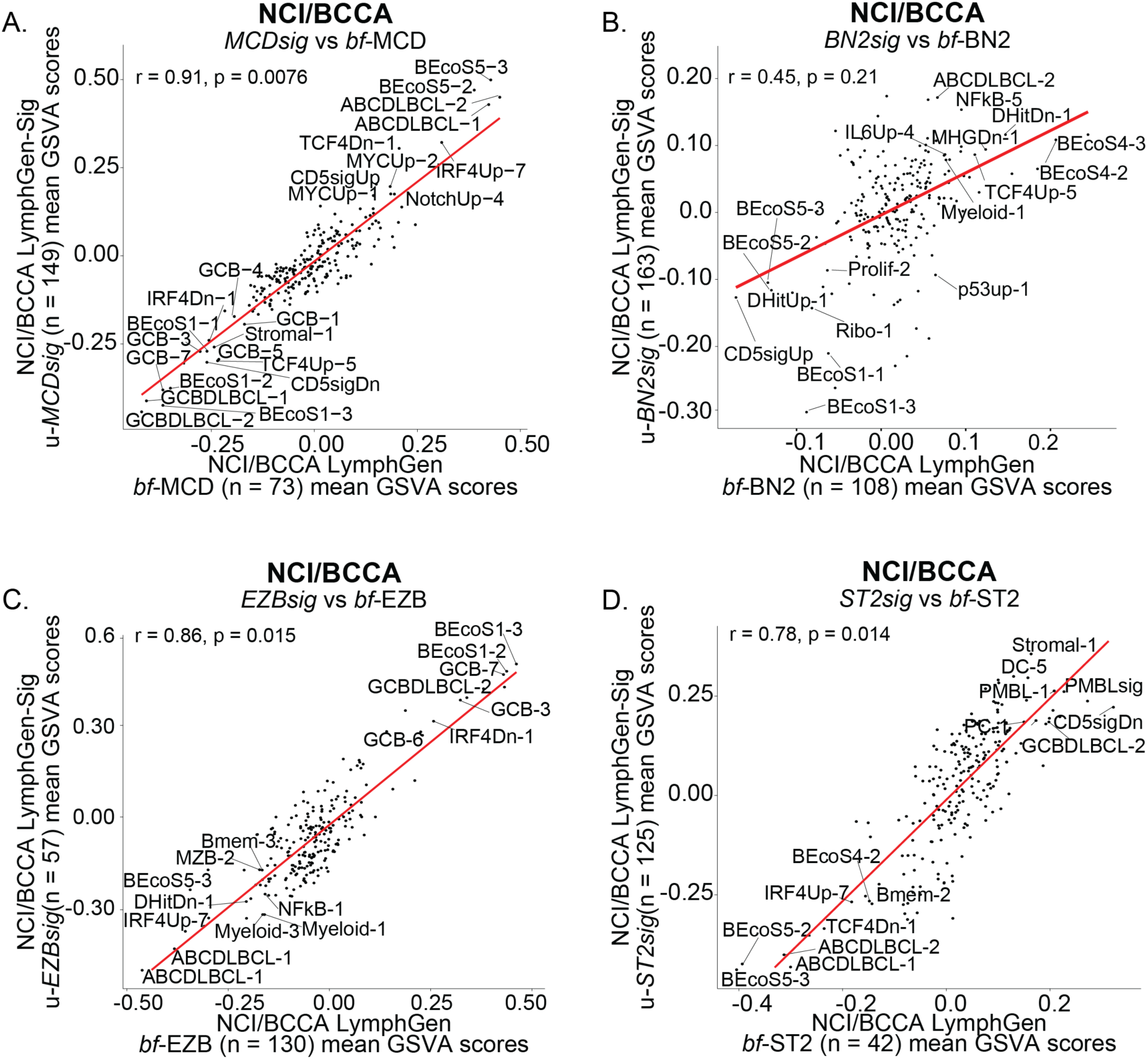
LG-unclassified DLBCLs are assigned into LGsig subtypes based on shared transcriptional features. **A–D.** Scatter plots showing the mean GSVA scores for selected lymphoma-related gene sets for bona fide (*bf*) LymphGen subtypes versus corresponding LymphGen-unclassified DLBCLs assigned to the same LGsig cluster in the NCI/BCCA dataset. Shown are comparisons of *bf*-MCD DLBCLs (x-axis) versus u-*MCDsig* DLBCLs (y-axis) (A), *bf*-BN2 versus u-*BN2sig* (B), *bf*-EZB versus u-*EZBsig* (C), and *bf*-ST2 versus u-*ST2sig* (D). P-values for the scatter plots were obtained using a permutation test for correlation, in which sample labels were randomly shuffled 1,000 times to generate a null distribution.

**Supplementary Figure 4.**
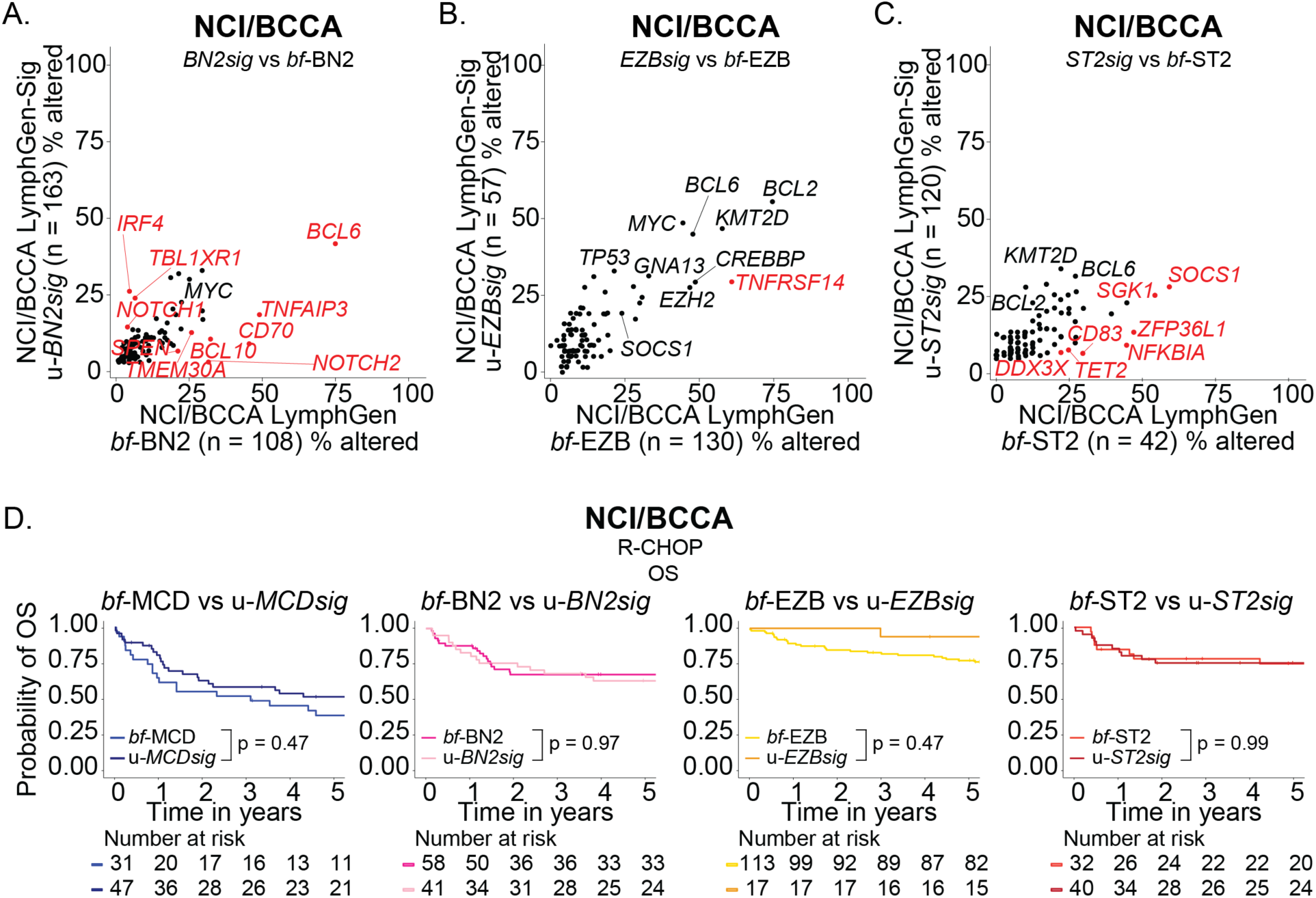
LG-unclassified DLBCLs are assigned into LGsig subtypes based on shared transcriptional features. **A–C.** Scatter plots showing the proportion of cases with genomic alterations in individual genes in LGsig subtypes (excluding bf-LG classifiable DLBCLs) compared with their corresponding *bf*-LymphGen genetic classes in the NCI/BCCA cohort: *BN2sig* vs *bf*-BN2 (A), *EZBsig* vs *bf*-EZB (B), *ST2sig* vs *bf*-ST2 (C). Dots highlighted in red are significantly enriched (Fisher’s exact test adj p < 0.1). **D.** Kaplan–Meier curves showing overall survival (OS) for R-CHOP–treated patients stratified by LymphGen genetic class and the corresponding LGsig transcriptional class in the NCI/BCCA cohort. P-values indicate comparisons between each LGsig group and its corresponding genetic subtype.

**Supplementary Figure 5.**
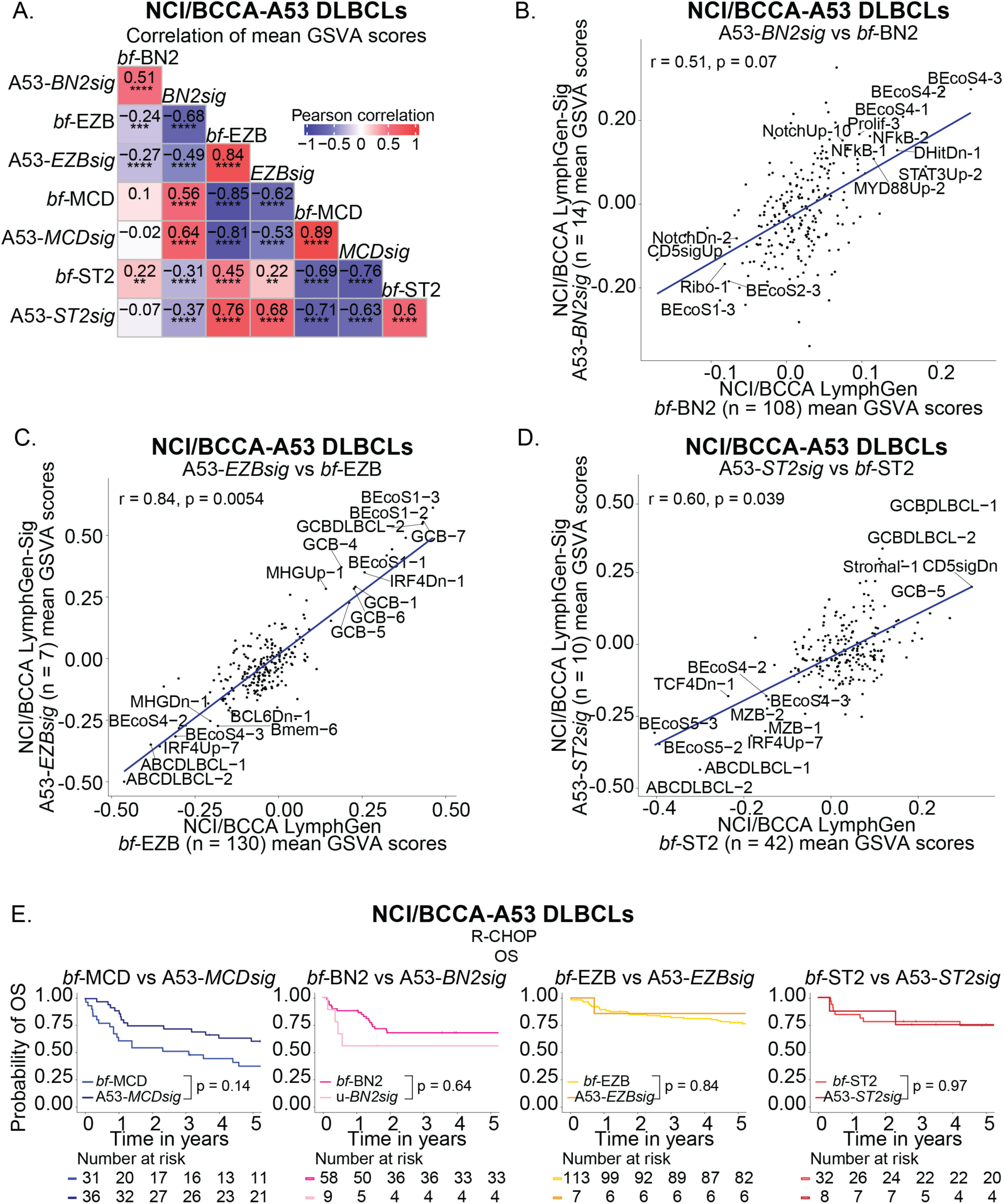
LGsig reassigns A53 LG DLBCLs. **A.** Correlation matrix showing pairwise correlations of mean GSVA scores between LymphGen genetic classes and A53-DLBCLs reassigned to LGsig transcriptional classes in the NCI/BCCA cohort. P-values were computed using a Pearson correlation t-test and adjusted for multiple testing using the Benjamini-Hochberg procedure. Adjusted p-values are shown (* adj p < 0.05, ** adj p < 0.01, ***adj p < 0.001, **** adj p < 0.0001). **B-D.** Scatter plots showing the mean GSVA scores for selected lymphoma-related gene sets for bona fide (*bf*) LymphGen subtypes versus the corresponding A53 DLBCLs reassigned to an LGsig cluster in the NCI/BCCA dataset. Shown are comparisons of, *bf*-BN2 versus A53-*BN2sig* (B), *bf*-EZB versus A53-*EZBsig* (C), and *bf*-ST2 versus A53-*ST2sig* (D). P-values for the scatter plots were obtained using a permutation test for correlation, in which sample labels were randomly shuffled 1,000 times to generate an empirical null distribution. **E.** Kaplan–Meier curves showing overall survival (OS) for R-CHOP–treated patients stratified by LymphGen genetic class and reclassified A53 DLBCLs NCI/BCCA cohort. P-values indicate comparisons between each A53-LGsig group and its corresponding LG genetic subtype.

**Supplementary Figure 6.**
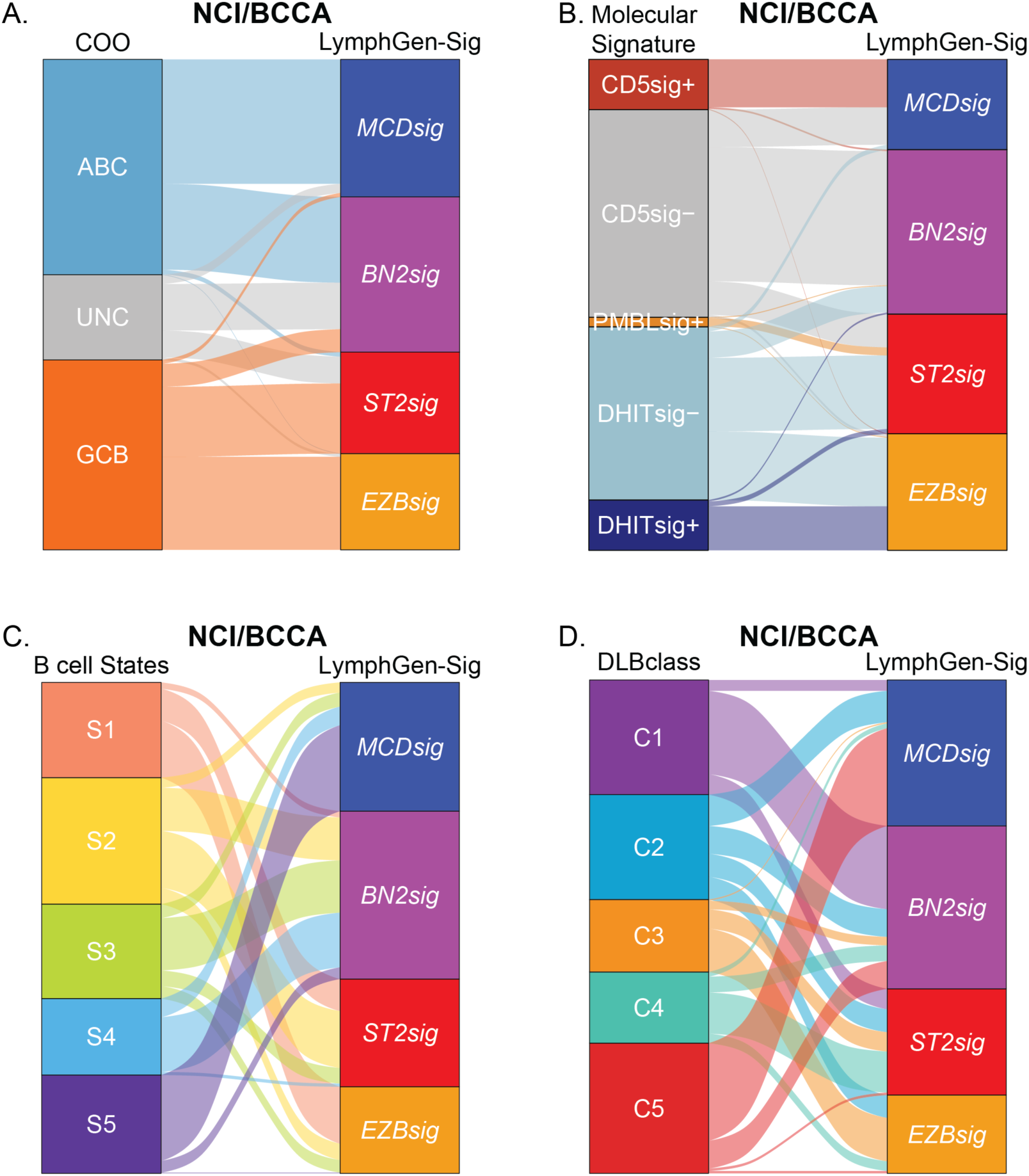
Comparison of LGsig to other molecular DLBCL classifiers. **A.** Alluvial plot showing the relationship between LGsig classes and cell-of-origin (COO) categories (ABC, UNC, and GCB). **B.** Alluvial plot showing the relationship between LGsig classes and selected molecular signatures, including CD5sig, PMBLsig, and DHITsig. **C.** Alluvial plot showing the relationship between LGsig classes and Ecotyper B-cell state signatures (S1–S5). **D.** Alluvial plot showing the relationship between LGsig classes and DLB*class* categories (C1–C5).

**Supplementary Figure 7.**
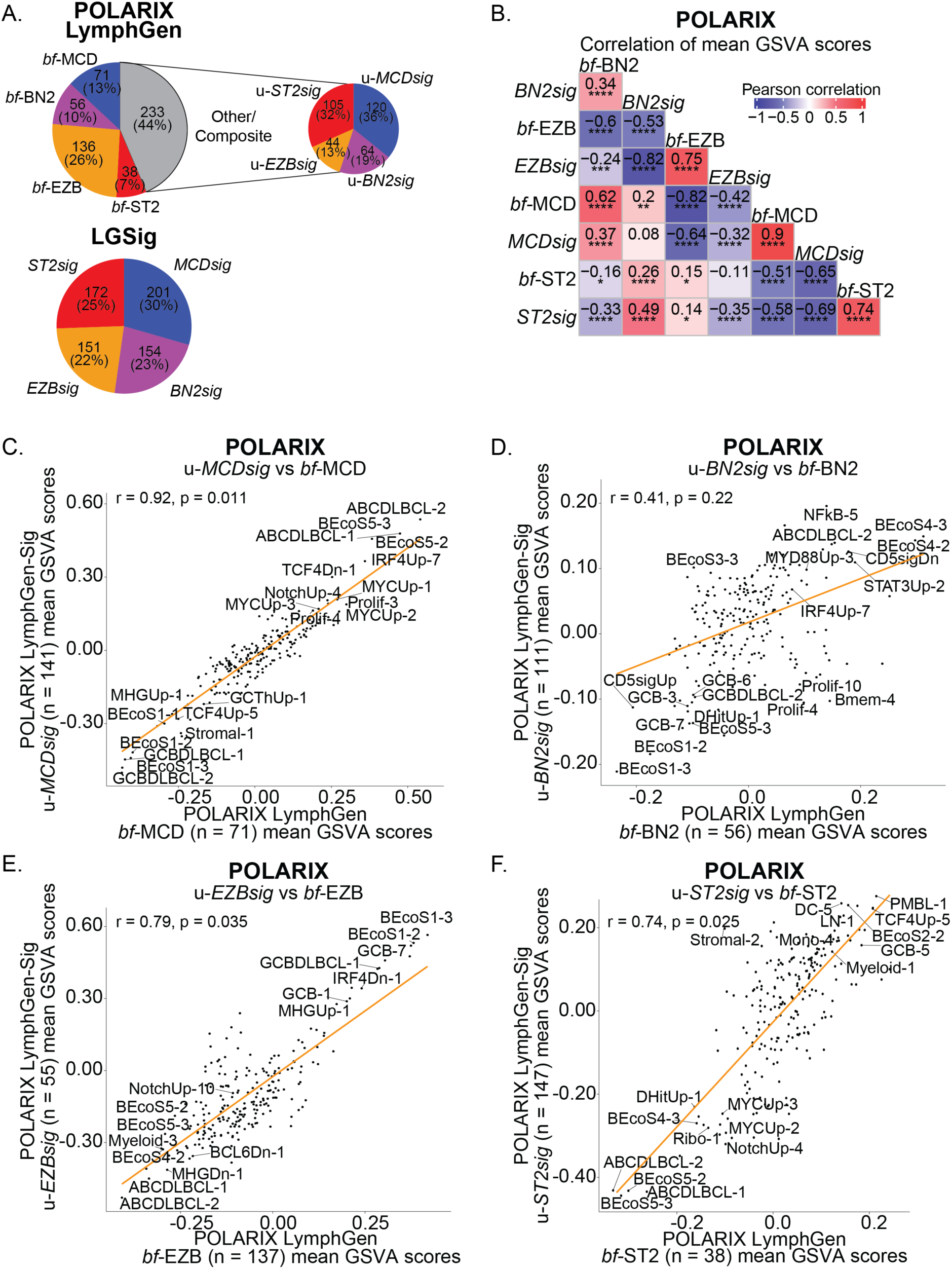
LGsig identifies LG-unclassified DLBCLs that benefit from pola-based chemoimmunotherapy in POLARIX. **A.** Top left: Pie chart showing LymphGen assignments for POLARIX cases, showing the proportion of *bf*-MCD, *bf*-BN2, *bf*-EZB, *bf-*ST2, and Other/Composite DLBCLs. Top right: subset of DLBCLs classified as Other/Composite by LymphGen, re-assigned using LGsig into *u*-MCDsig, *u*-BN2sig, *u*-EZBsig, or *u*-ST2sig. Bottom: overall LGsig subtype distribution across the POLARIX cohort. Numbers indicate sample counts and percentages are shown in parentheses. **B.** Correlation matrix showing pairwise correlations of mean GSVA scores between LymphGen genetic classes and LGsig transcriptional classes in the POLARIX cohort. P-values were computed using a Pearson correlation t-test and adjusted for multiple testing using the Benjamini-Hochberg procedure. Adjusted p-values are shown (* adj p < 0.05, ** adj p < 0.01, ***adj p < 0.001, **** adj p < 0.0001). **C–E.** Scatter plots showing the mean GSVA scores for selected lymphoma-related gene sets for bona fide (*bf*) LymphGen subtypes versus the corresponding LymphGen-unclassified DLBCLs assigned to an LGsig cluster in the POLARIX cohort. Shown are comparisons of *bf*-MCD DLBCLs (x-axis) versus u-*MCDsig* DLBCLs (y-axis) (B), *bf*-BN2 versus u-*BN2sig* (C), *bf*-EZB versus u-*EZBsig* (D), and *bf*-ST2 versus u-*ST2sig* (E).

**Supplementary Figure 8.**
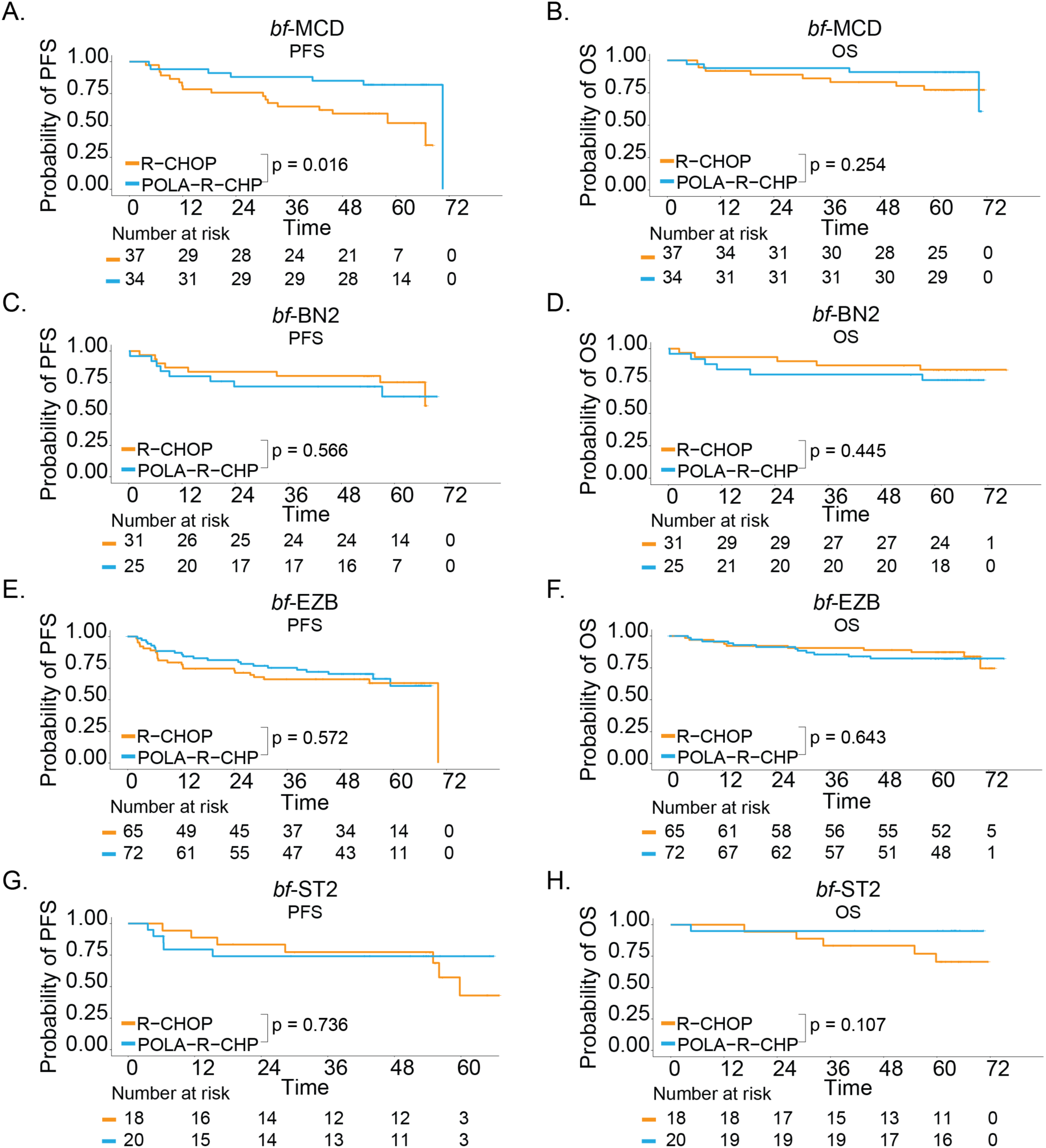
LGsig identifies LG-unclassified DLBCLs that benefit from pola-based chemoimmunotherapy in POLARIX. **A–B**. Kaplan–Meier curves comparing progression-free survival (A) and overall survival (B) between R-CHOP- and pola–R–CHP-treated *bf*-MCD DLBCLs in the POLARIX cohort. **C–D.** Kaplan–Meier curves comparing progression-free survival (C) and overall survival (D) between R-CHOP- and pola–R–CHP-treated *bf*-BN2 DLBCLs in the POLARIX cohort. **E–F**. Kaplan–Meier curves comparing progression-free survival (E) and overall survival (F) between R-CHOP- and pola–R–CHP-treated *bf*-EZB DLBCLs in the POLARIX cohort. **G–H**. Kaplan–Meier curves comparing progression-free survival (G) and overall survival (H) between R-CHOP- and pola–R–CHP-treated *bf*-ST2 DLBCLs in the POLARIX cohort. Numbers at risk are shown below each plot. Unadjusted p-values are displayed.

**Supplementary Figure 9.**
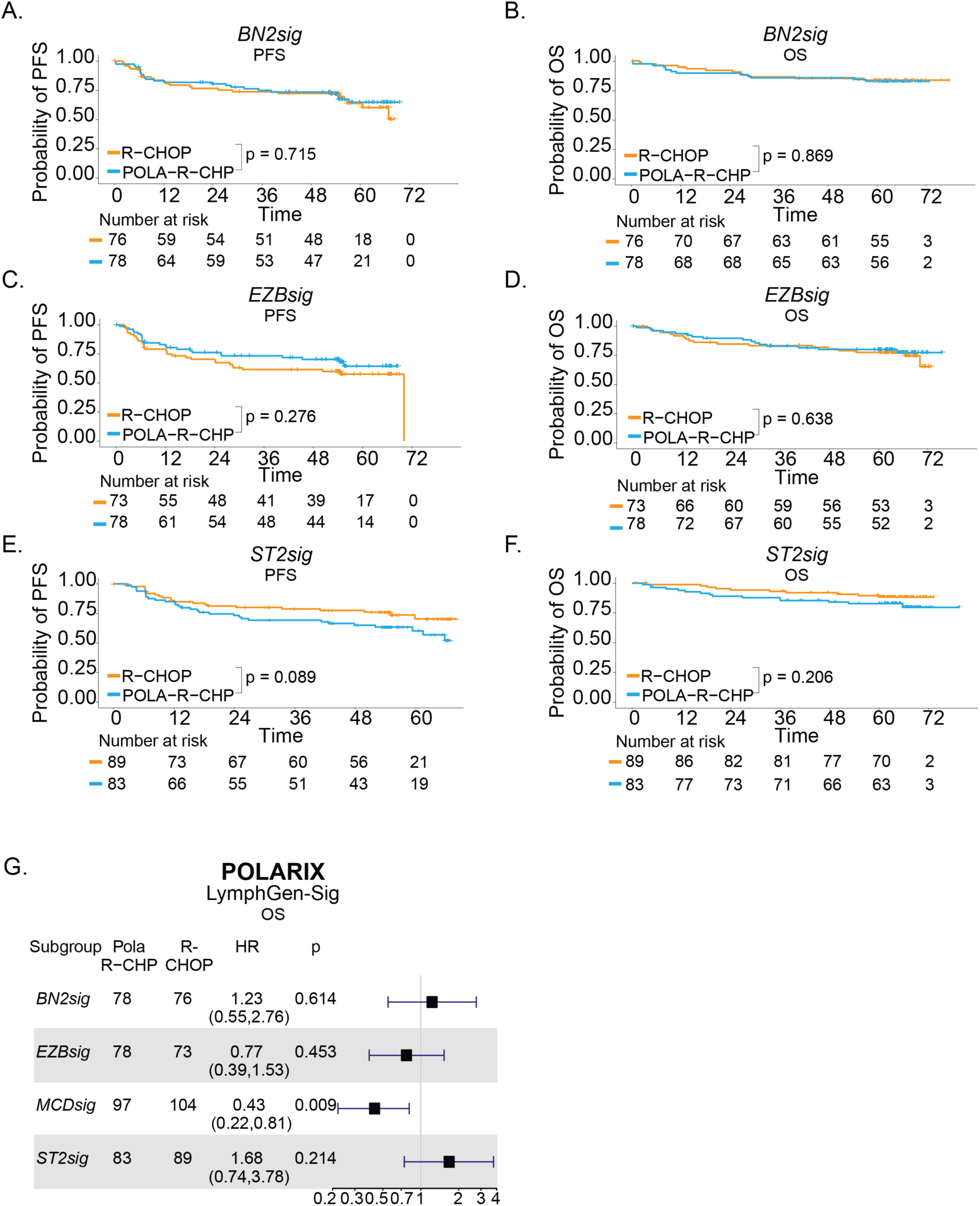
LGsig identifies LG-unclassified DLBCLs that benefit from polabased chemoimmunotherapy in POLARIX. **A–B.** Kaplan–Meier curves comparing progression-free survival (A) and overall survival (B) between R-CHOP- and pola–R–CHP-treated *BN2sig* DLBCLs in the POLARIX cohort. **C–D**. Kaplan–Meier curves comparing progression-free survival (C) and overall survival (D) between R-CHOP- and pola–R–CHP-treated *EZBsig* DLBCLs in the POLARIX cohort. **E–F**. Kaplan–Meier curves comparing progression-free survival (E) and overall survival (F) between R-CHOP- and pola–R–CHP-treated *ST2sig* DLBCLs in the POLARIX cohort. Numbers at risk are shown below each plot. Unadjusted p-values are displayed. **G.** Forest plot showing overall survival hazard ratios for pola–R–CHP versus R-CHOP across LGsig subgroups. Hazard ratios are adjusted for International Prognostic Index score (2 vs 3–5), age (<60 vs ≥60 years), and COO (activated B cell, germinal center B cell, unclassified, unknown).

**Supplementary Figure 10.**
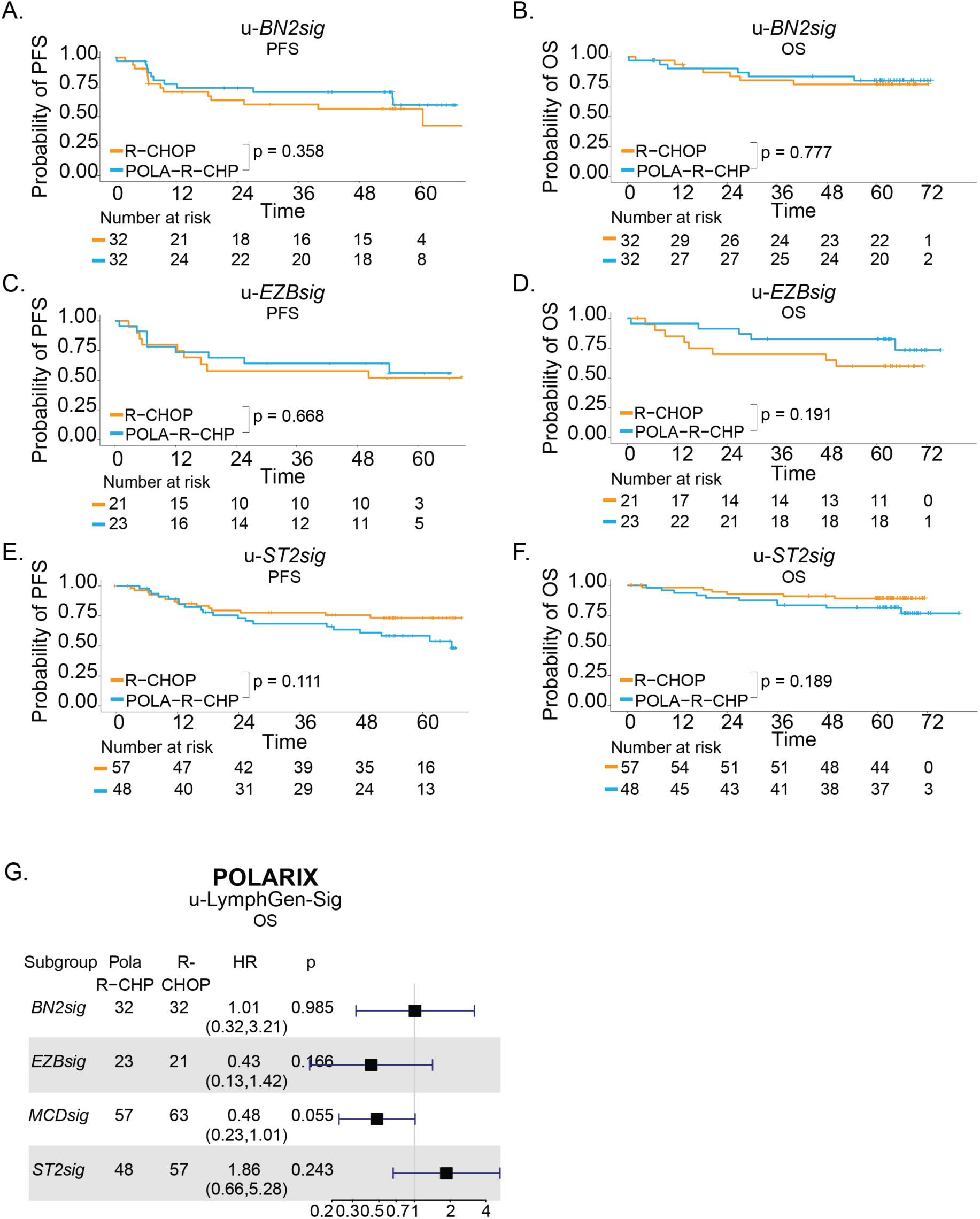
LGsig identifies LG-unclassified DLBCLs that benefit from polabased chemoimmunotherapy in POLARIX. **A–B**. Kaplan–Meier curves comparing progression-free survival (A) and overall survival (B) between R-CHOP- and pola–R–CHP-treated u-*BN2sig* DLBCLs in the POLARIX cohort. **C–D**. Kaplan–Meier curves comparing progression-free survival (C) and overall survival (D) between R-CHOP- and pola–R–CHP-treated u-*EZBsig* DLBCLs in the POLARIX cohort. **E–F**. Kaplan–Meier curves comparing progression-free survival (E) and overall survival (F) between R-CHOP- and pola–R–CHP-treated u-*ST2sig* DLBCLs in the POLARIX cohort. Numbers at risk are shown below each plot. Unadjusted p-values are displayed. **G.** Forest plot showing overall survival (OS) hazard ratios for pola–R–CHP versus R-CHOP across u-LGsig subgroups. Hazard ratios are adjusted for International Prognostic Index score (2 vs 3–5), age (<60 vs ≥60 years), and COO (activated B cell, germinal center B cell, unclassified, unknown).

