## Supplemental Data 1 for "LymphGen-Sig: Integrating genetic and transcriptional states to predict therapeutic response in DLBCL"

**Supplemental Methods**

**Comparison of transcriptomes between groups**

Gene sets were curated from the Staudt lab’s Signature database (SigDB)^1^. Gene sets related to lymphoma, normal immune cell populations, and relevant oncogenic processes that passed a size filter of greater than or equal to 10 genes and less than or equal to 500 genes were kept; this left 220 gene sets. Additional gene sets related to molecular DLBCL classifications were included, for a total of 223 gene sets^2,3^. For each gene set, the expression of its constitutive genes was quantified into a score using Gene Set Variation Analysis (gsva package in R (v1.4.6.0))^4^. To compare two groups, GSVA score for each gene set was calculated for each sample in each group, and the mean scores per gene set per group were compared in a scatter plot with the Pearson correlation coefficient. Significance was determined through a permutation test: the correlation was recomputed for each of 1000 iterations where the sample labels were shuffled and the mean expression scores were recalculated. A normal approximation of the distribution of correlations was used to calculate the right-tailed p-value for the unpermuted correlation.

**Gene alteration analysis**

*LymphGen classifications:* LymphGen v2.0 was run using the web-based portal (<https://llmpp.nih.gov/lymphgen/index.php>). Samples with a probability of subtype assignment of greater than or equal to 90% were defined as “core” samples, and samples with a probability of subtype assignment of greater than or equal to 50% were defined as “extended” samples, as previously described ^5^. LymphGen was run on samples from each cohort independently.

*DLBclass classifications*: DLBclass classifications for available NCI samples were downloaded from Chapuy et al. 2025^6^.

**Comparison of genetic alterations between groups**

Analysis of genetic alterations was restricted to DLBCL driver genes captured in the BCCA targeted sequencing panels^7,8^ and LymphGen hallmark alterations. For each comparison, genes were excluded if they were not altered at least 5 cases in at least one of the groups being compared.
